# Acquisition of Group B streptococcus colonization in preterm pregnancy

**DOI:** 10.64898/2026.08.21.26361025

**Authors:** Aaron Bowers, John Elliott, Nicole Book, Sanchita Krishna, Emily Hamburg-Shields

## Abstract

**Objective:** The purpose of this study was to estimate the negative predictive value (NPV) of screening for group B streptococcus (GBS) colonization in pregnant patients undergoing antepartum hospitalization for GBS colonization status at the time of preterm delivery.

**Study Design:** This prospective, observational cohort study compared GBS colonization status upon initial hospital admission to that at the time of delivery. Pregnant patients at 22 to 35 weeks gestation admitted to the antepartum unit at a tertiary care hospital underwent standard screening for GBS colonization. When preterm labor progressed or iatrogenic preterm delivery was indicated, the GBS colonization test was repeated. Comparison of the sequential test results was performed to determine the NPV of the antepartum screening test for the intrapartum status.

**Results:** 159 eligible patients were enrolled in the study, and 100 completed the study and were included in the analysis. The average gestational age at admission was 30 weeks 1 day (95% confidence interval [CI] 29w4d to 30w6d) and the average duration of pregnancy latency in the study group was 17.5 days (95% CI 15.1 to 19.8). GBS colonization rate at the time of admission was 18% and at the time of delivery was 20%. The NPV of GBS screening at admission was 91.5% (95% CI 83.2 to 96.5%) and the positive predictive value (PPV) was 72.2% (95% CI 46.4 to 90.3%).

**Conclusion:** In a cohort of pregnant patients with preterm pregnancy complications, GBS screening at the time of antepartum hospital admission has an NPV of 91.5% (95% CI 83.2 to 96.5%) for GBS colonization at the time of preterm delivery. This is comparable to the published NPV of routine GBS screening for colonization status at term delivery.

**Key Points:**

- Prevalence of GBS colonization was 18% at preterm hospital admission and 20% at preterm delivery.
- The NPV of initial GBS screening for status at preterm delivery was 91.5%.
- 7 of 100 patients had false-negative GBS screening.

## Introduction

Neonatal early-onset group B streptococcal (GBS) disease (EOGBD) causes sepsis, pneumonia, and meningitis in the first week of life following exposure to commensal GBS from the genital tract of the colonized parturient^1^. To detect pregnancies at risk for neonatal EOGBD, universal antenatal screening for GBS colonization is routinely performed between 36 and 38 weeks gestation in the United States^2,3^. The rationale for the timing of routine screening is the transient nature of GBS colonization in pregnancy, therefore the predictive value of screening at an earlier gestational age is lower than when it is performed at or near term^4–7^. Between 10 and 30% of pregnant people exhibit GBS colonization during routine third trimester screening. Previous studies in cohorts of term or near-term patients have shown a positive predictive value of 55% (95% CI 46-63%) and negative predictive value of 98% (95% CI 96-99%) for GBS colonization status at term delivery^2,8–10^. Patients with a positive screening result are treated during labor with intrapartum antibiotic prophylaxis (IAP). Patients undergoing cesarean delivery receive preoperative antibiotics regardless of GBS colonization. GBS screening and IAP represent a successful public health accomplishment that has substantially reduced the rate of vertical transmission of GBS and subsequent neonatal morbidity and mortality due to EOGBD^11–14^.

In spite of the success of universal antenatal screening and intrapartum antibiotic prophylaxis, GBS remains the leading cause of early-onset neonatal sepsis, pneumonia and meningitis in the United States (0.23 cases per 1000 live births)^15,16^. One of the factors that contributes to the continued incidence of invasive GBS disease is the elevated susceptibility of preterm neonates, a population that comprises 10.4% of live births in the United States^17^. Even with adherence to the current screening and prophylaxis guidelines, preterm infants are 4 times more likely than term infants to develop EOGBD^12^, and have increased risk of death from EOGBD (19.2% case fatality rate in preterm infants compared to 2.1% in term infants)^16^.

Pregnant patients undergoing antepartum hospital admission prior to anticipated preterm delivery due to complications such as preterm labor, preterm premature rupture of membranes (PPROM), and preterm preeclampsia typically have an unknown GBS colonization status at the time of hospital admission. The standard of care in this scenario is to screen for GBS colonization at the time of admission to the hospital. If the patient is in active preterm labor with unknown GBS colonization status, empiric IAP is administered until negative GBS colonization is demonstrated or until preterm labor halts. If the pregnancy continues and the initial screening test was negative, then GBS status is re-screened after 5 weeks due to risk of acquired colonization^13,14,18^. This 5-week interval is a recommendation from the American College of Obstetrics & Gynecology (ACOG) which has been endorsed by the American Academy of Pediatrics and the Society for Maternal Medicine^19^. To our knowledge, there are not any published studies that have examined the stability of GBS colonization over this 5-week interval at preterm gestational ages, even though it is well-established that GBS colonization is transient and variable during normal pregnancy^18^. We sought to address this gap by determining the predictive value of GBS screening upon antepartum admission for colonization status at the time of preterm delivery.

We hypothesized that GBS screening in hospitalized gravidas at preterm gestational ages would have an inferior negative predictive value (NPV) for GBS colonization status at the time of preterm delivery compared to the established NPV for routine GBS screening in patients with uncomplicated pregnancies. This hypothesis was formulated to address whether an antepartum GBS screening result is valid for the recommended 5-week duration in a population at elevated risk for EOGBD.

## Methods

This prospective, observational cohort study was carried out in the obstetric unit of a single tertiary care center with approval and supervision of the Institutional Review Board (IRB) of the OhioHealth Research Institute. Patients were recruited beginning in August 2018 until the institution’s research activity was paused in March 2020 due to safety considerations during the COVID-19 pandemic. Recruitment was resumed in December 2021 after receiving renewed IRB approval and completed in September 2023 after achievement of the target sample size.

Our study population consisted of pregnant patients admitted to the antepartum unit between 22 weeks 0 days and 34 weeks 6 days of gestation who underwent an initial GBS screening test and were anticipated to remain admitted until preterm delivery. Patients with a pre-existing indication for IAP (GBS bacteriuria or history of EOGBD in a prior neonate) did not require screening and thus were excluded from the study. All patients received antibiotics as clinically indicated, typically IAP for unknown GBS status in preterm labor, or latency antibiotics for preterm premature rupture of membranes (PPROM). The upper gestational age limit of 34 weeks 6 days was chosen because at the initiation of the study, routine near-term screening for GBS colonization was recommended to occur as early as 35 weeks gestation^20^. To decrease the possibility of a false negative screening test, patients were not enrolled if they had received antibiotics within the past 24 hours. To participate in the study, patients were required to be at least 18 years of age and able to ready study documents and provide informed consent.

We estimated that 100 subjects were needed to detect a clinically relevant difference in the primary outcome, NPV of GBS screening for preterm delivery, compared to the published NPV of routine near-term GBS screening in the general population. In a high-quality published study, the NPV of routine screening for GBS colonization at term was determined to be 95% with a CI of 93-96.5%^21^. With an assumed positive GBS screening result of 25% (based on the known prevalence of GBS colonization in the obstetric population)^19^, we determined that a sample size of 100 patients would detect 10% or greater absolute deficit in NPV of GBS screening in preterm pregnancy compared to the known NPV of GBS screening for term delivery.

Upon admission to labor and delivery or the antepartum unit, patients at preterm gestational age underwent an initial GBS screening test by clinical staff as clinically indicated. If a patient was receiving parenteral antibiotics at the time of admission (for example, IAP initiated at a referring hospital before transfer to the admitting antepartum unit), the screening test was not performed until 24 hours after antibiotic therapy ceased, per unit protocol. Study personnel reviewed the obstetric patient census to identify eligible patients, and those who met inclusion criteria were enrolled after giving their informed consent. Clinical and demographic data were recorded in a secure REDCap database hosted by the OhioHealth Research Institute^22^. In enrolled patients at the time of spontaneous preterm labor or the decision to proceed with iatrogenic preterm delivery at 36w6d or earlier, a repeat GBS test was performed by clinical staff. For both screening and experimental GBS testing, samples were obtained by vaginoperineal or vaginorectal swab and were incubated in an enrichment broth for 18-24 hours and the presence of Group B streptococcus isolate was evaluated by polymerase chain reaction per the institutional laboratory protocol. Study sample collection was not performed in patients who had received parenteral antibiotics within the previous 24 hours or in the event of clinical urgency or emergency. Patients who achieved gestational age 37 weeks or greater dropped out of the study.

Descriptive information on the study sample was tabulated. Means and standard deviations were used for continuous variables and percentages used for nominal (categorical) variables.

Tabulations of GBS initial screening results and the repeat GBS test at the time of delivery were conducted along with 95% CI for sensitivity, specificity, PPV and NPV. All analyses conducted with SPSS version 25 and Vassar Clinical Calculator^23^. The STROBE checklist for observational cohort studies was used as a guide for this manuscript (Supplement)^24^.

## Results

In 41 months of active study recruitment, 159 patients were enrolled to achieve 100 completed study cases (Figure 1). The demographics and characteristics of the study cohort are summarized in Table 1. The study population had a mean age of 30.7 years (interquartile range 24.7 to 36.7 years) and was predominantly nulliparous (56%). Of the 44 multiparous patients in our study, 18 (40.9%) had a history of preterm delivery.

**Figure 1.**
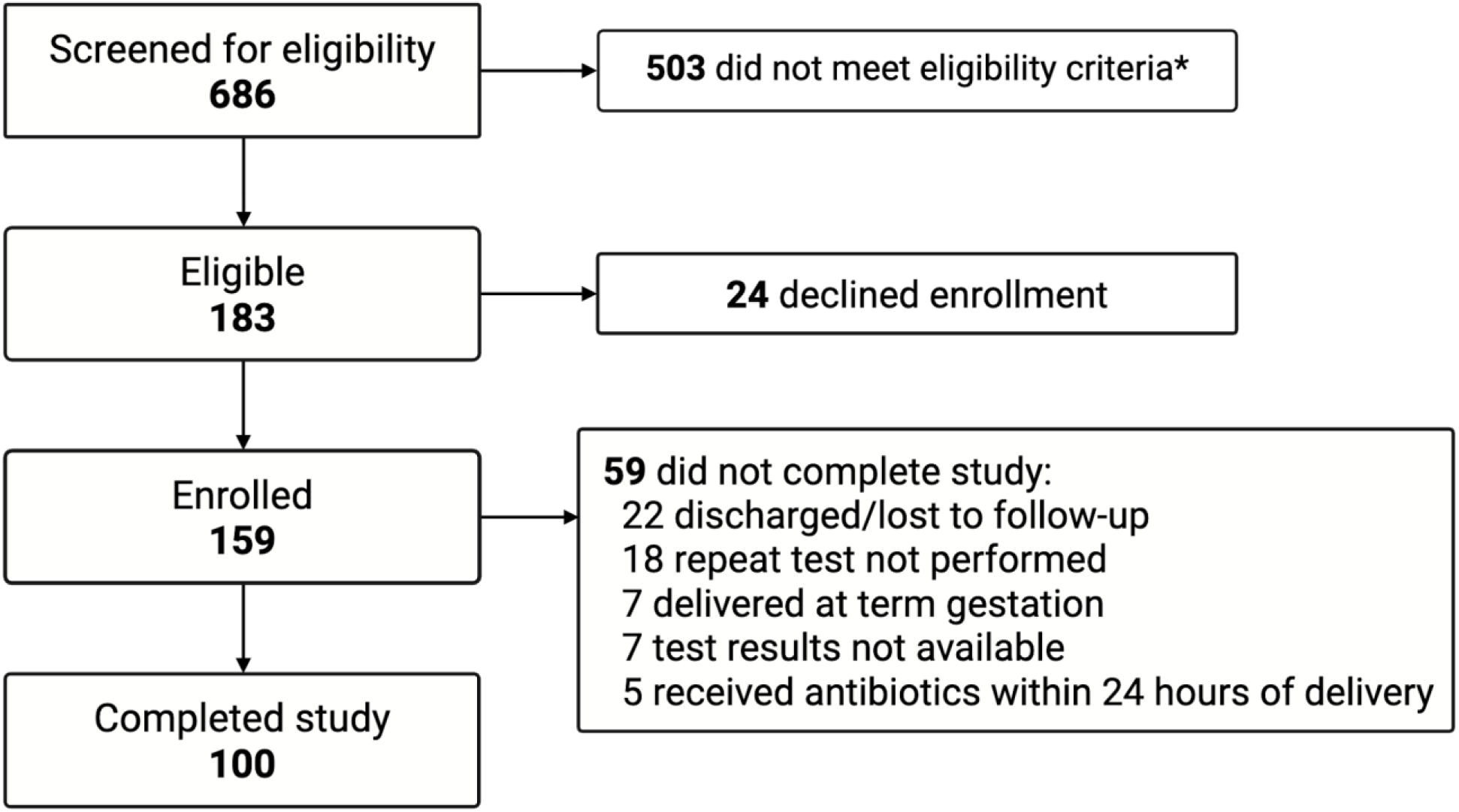
STROBE flow sheet of study enrollment. *Most common reasons for exclusion were pregnancies outside of gestational age criteria, non-English-speaking and low clinical suspicion for risk of preterm delivery. Figure created using BioRender.

**Table 1.**
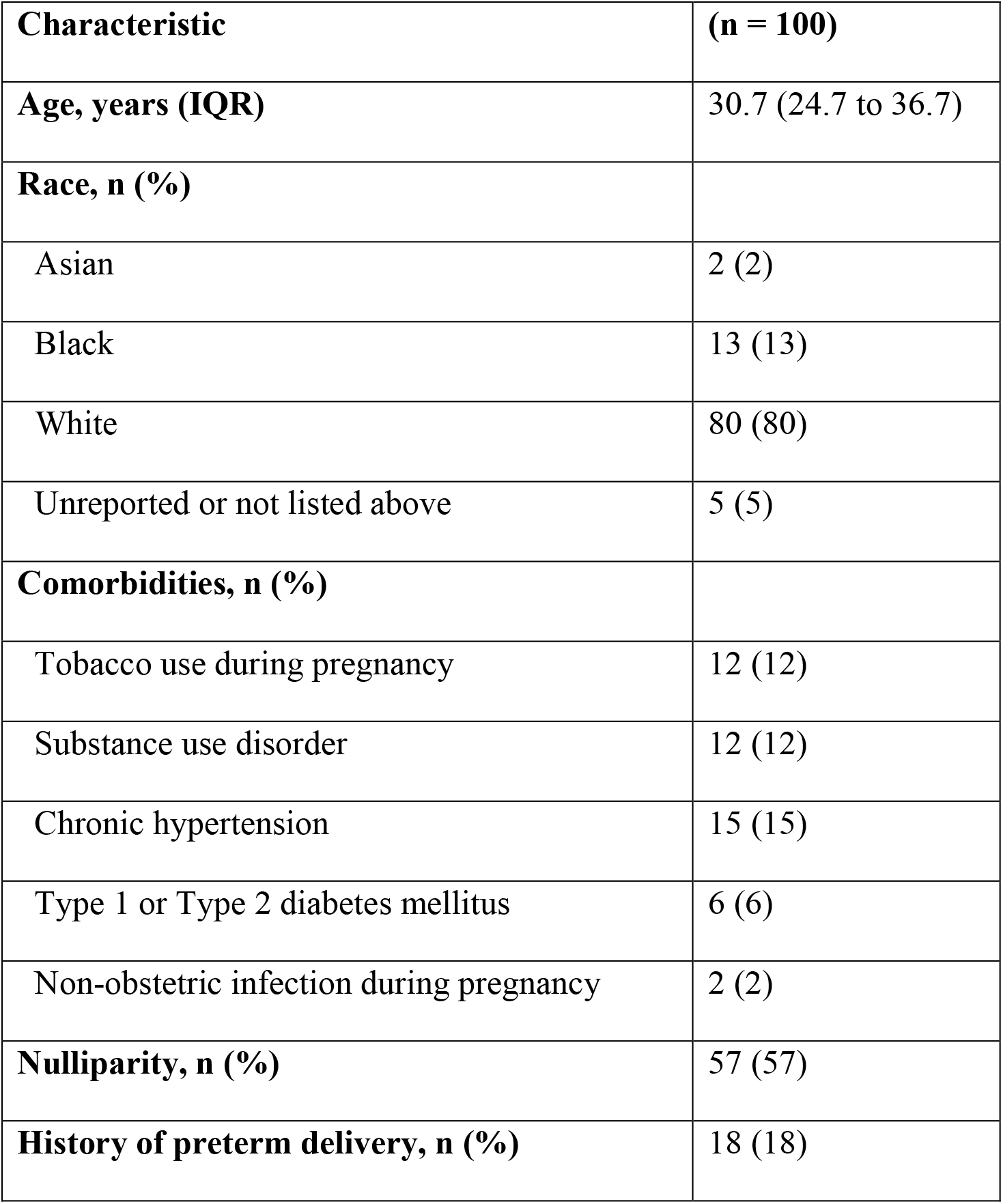
Patient characteristics.

| **Characteristic** | **(n = 100)** |
| --- | --- |
| **Age, years (IQR)** | 30.7 (24.7 to 36.7) |
| **Race, n (%)** |  |
| Asian | 2 (2) |
| Black | 13 (13) |
| White | 80 (80) |
| Unreported or not listed above | 5 (5) |
| **Comorbidities, n (%)** |  |
| Tobacco use during pregnancy | 12 (12) |
| Substance use disorder | 12 (12) |
| Chronic hypertension | 15 (15) |
| Type 1 or Type 2 diabetes mellitus | 6 (6) |
| Non-obstetric infection during pregnancy | 2 (2) |
| **Nulliparity, n (%)** | 57 (57) |
| **History of preterm delivery, n (%)** | 18 (18) |

Common indications for hospital admission in the study cohort included hypertensive disorders of pregnancy (62%), PPROM (31%), and preterm labor/advanced cervical dilation (25%). Many patients had more than one obstetric complication (Table 2). The average gestational age at admission was 30 weeks 1 day (95% CI 29w4d to 30w6d). The average interval between the screening test and intrapartum test was 17.5 days (SD 11.9 days) and the average gestational age at the time of delivery was 32 weeks 5 days (SD 22 days).

**Table 2.** Pregnancy complications and outcomes.

| **Characteristic** | **(n = 100)** |
| --- | --- |
| **Gestational age at admission, mean (range)** | 30w1d (22w5d to 34w6d) |
| **Gestational age at delivery, mean (range)** | 32w5d (25w1d to 36w6d) |
| **Complications, n (%)** |  |
| Preterm premature rupture of membranes | 31 (31) |
| Preterm labor/Advanced cervical dilation | 25 (25) |
| Preeclampsia/Gestational hypertension | 62 (62) |
| Intrauterine growth restriction | 17 (17) |
| Abnormal placentation (previa, accreta) | 9 (9) |
| Placental abruption | 11 (11) |
| Multiple gestation | 15 (15) |
| **Type of labor, n (%)** |  |
| Spontaneous labor | 20 (20) |
| Induced labor | 33 (33) |
| None (pre-labor cesarean delivery) | 47 (47) |
| **Mode of delivery, n (%)** |  |
| Vaginal and operative delivery | 36 (36) |
| Cesarean delivery | 64 (64) |

Eighteen patients (18%) had a positive GBS screening test on admission to the hospital, and 20 patients (20%) had a positive test at the time of delivery (Table 3). Therefore, in this cohort, the admission GBS screening had a sensitivity of 65% (95% CI 40.8 to 84.6%) and specificity of 93.8% (95% CI 86 to 97.9%) for GBS colonization at the time of delivery. The PPV was 72.2% (95% CI 46.4 to 90.3%) and the NPV was 91.5% (95% CI 83.2 to 96.5%). This NPV is comparable to the published NPV of 95% (95% CI 93% to 96.5%)^21^. Seven cases had a false negative screening test at hospital admission with positive GBS colonization at the time of delivery; the interval between the two tests in these cases ranged from 7 to 36 days (Table 4).

**Table 3.** 2-by-2 table constructed for calculation of sensitivity, specificity, positive predictive value, and negative predictive value of admission GBS screening test with 95% confidence intervals (CI) reported.

|  |  | **Intrapartum GBS test result** |  |
| --- | --- | --- | --- |
|  |  | Positive | Negative |
| **Admission GBS screening result** | Positive | 13 | 5 |
|  | Negative | 7 | 75 |
| Sensitivity: 65% (95% CI 40.8 to 84.6%)<br>Specificity: 93.8% (95% CI 86 to 97.9%)<br>PPV: 72.2% (95% CI 46.4 to 90.3%)<br>NPV: 91.5% (95% CI 83.2 to 96.5%) |  |  |  |

**Table 4.** Characteristics of cases with conversion from GBS negative to GBS positive status.

|  | Gestational age at admission | Admission Diagnosis | Number of days between tests | Antibiotics received during admission |
| --- | --- | --- | --- | --- |
| Patient 1 | 24w2d | Placental abruption | 14 | None |
| Patient 2 | 31w1d | Preeclampsia with severe features | 36 | None |
| Patient 3 | 32w1d | Preeclampsia with severe features | 7 | None |
| Patient 4 | 29w1d | PPROM | 19 | Latency antibiotics<sup>a</sup> |
| Patient 5 | 28w5d | PPROM | 36 | Latency antibiotics<sup>a</sup> |
| Patient 6 | 23w0d | PPROM | 15 | Latency antibiotics<sup>a</sup> |
| Patient 7 | 33w2d | Placenta previa | 22 | None |

<sup>a</sup>For management of preterm prelabor rupture of membranes (PPROM), patients 4 and 5 received 2g of intravenous ampicillin every 6 hours and 500mg intravenous azithromycin daily for 2 days followed by 500mg of oral amoxicillin every 8 hours and oral 250mg azithromycin daily for 5 days. Patient 6 was treated with a modified regimen of vancomycin and azithromycin due to drug allergies.

## Discussion

Multiple studies in near-term and term pregnancy have demonstrated high negative predictive values of GBS screening within 5 weeks prior to delivery for GBS colonization status in labor— ranging from 87.2 to 97.1% across multiple studies^25,26^. These findings have served as the basis for screening 5 weeks in advance of anticipated delivery. The recommended gestational age window for screening is between 36 and 38 weeks gestation to allow time for culture results to be available before the typical onset of spontaneous labor or the timing of indicated delivery^19^. The aim of this study was to calculate the NPV of GBS screening at the time of admission for preterm delivery. The NPV is the outcome of interest because, for the purposes of identifying candidates for IAP to prevent EOGBD, it is most valuable to have a test with a low false-negative rate. In the case of low-risk interventions such as IAP, false positive tests are generally considered to be more tolerable than false negatives^27^. Our study population included pregnant patients with diverse obstetric diagnoses and gestational ages. The enrolled patients had a high rate of preterm delivery, reflecting the intention of the study to focus on pregnancies at the highest risk of producing neonates that are vulnerable to EOGBD. The NPV of GBS screening in our population at the time of preterm delivery was 91.4%, which is comparable to the NPV of near-term screening in previously published studies^28,29^. This finding supports our null hypothesis.

The clinical approach to prevention of GBD in both term and preterm infants may be refined in the future by the implementation of GBS vaccination and rapid intrapartum screening tests^1^. Prenatal vaccination to confer neonatal immunity to GBS via transplacental antibodies is currently being developed^30,31^ and is anticipated to be cost-effective^32,33^ and to accomplish a significant advance beyond IAP by being able to prevent late-onset group B streptococcus disease. It will remain important to understand the dynamics of GBS colonization at preterm gestational ages, since several of the recommended maternal vaccinations for neonatal benefit are typically not administered until the third trimester in order to maximize the degree of passive immunity conferred to the infant^2^. Rapid intrapartum testing, which is based on nucleic acid detection without enrichment broth incubation, may be informative for guiding IAP and postnatal antibiotic prophylaxis in both term and preterm neonates^34^.

The generalizability of this study’s findings are limited in part because it was carried out at a single tertiary care center. Due to the observational nature of the study and the inclusion of all obstetric complications with anticipated preterm delivery, there was inherent variability in pregnancy latency and antibiotic exposure. Patients admitted with preterm labor and PPROM were under-represented in comparison to patients with iatrogenic preterm delivery; dynamics of GBS colonization may vary between these etiologies of preterm delivery, and this study did not have sufficient sample size to perform subgroup analyses by obstetric complication. Future studies to advance EOGBD prevention in preterm neonates may benefit from stratification by maternal obstetric complications and other specific neonatal risk factors such as degree of prematurity. This study’s finding of comparable NPV to that of routine GBS colonization screening in the general population supports that the increased rate of EOGBD in preterm neonates may be due to biological susceptibility, rather than a deficit in the predictive value of routine maternal screening at preterm gestational ages.

## Data Availability Statement

The underlying research data of this article are not publicly available due to patient privacy and small sample size. The data are available from the corresponding author on reasonable request.

## Acknowledgements

Audrey McDonald, CRC; Johnna Booker, CRC; Amanda Bliemeister, PhD, MSN, RN; Carla Castillo Payne, BSPH; and Christina Belcher, BSN, RN, CRC are acknowledged for their contribution to patient enrollment and clinical data abstraction. We also thank the enrolled patients and the house staff and antepartum nurses of Riverside Methodist Hospital for their support of this study.

## Declaration of Generative AI Use

The authors confirm that generative AI was not used in the preparation of this manuscript.

